# Platelet polyphosphate and SARS-Cov-2 mRNA-vaccine-induced inflammatory side effects: a pilot study

**DOI:** 10.1101/2021.09.13.21263437

**Authors:** Takashi Uematsu, Atsushi Sato, Hachidai Aizawa, Tetsuhiro Tsujino, Taisuke Watanabe, Kazushige Isobe, Hideo Kawabata, Yutaka Kitamura, Takaaki Tanaka, Tomoyuki Kawase

**Affiliations:** Tokyo Plastic Dental Society, Tokyo, Japan; Department of Materials, Science and Technology, Niigata University, Niigata, Japan; Division of Oral Bioengineering, Institute of Medicine and Dentistry, Niigata University, Niigata, Japan

**Keywords:** COVID-19, blood platelet, polyphosphates, sex, adverse effects

## Abstract

**Background:** Platelets have recently been recognized as immune cells. Platelets first contact invading pathogens and then induce immune reactions in cooperation with white blood cells. Platelet polyphosphate (polyP), which is classically recognized as a thrombotic and hemostatic biomolecule, has recently attracted attention as a ‘cytokine’ that modulates inflammation and is involved in intercellular communication between platelets and major immune cells.

**Objective:** To determine the involvement of polyP in SARS-Cov-2-mRNA vaccine-induced immune responses, this pilot study examined the effects of mRNA vaccines on platelet polyP levels.

**Methods:** Before and after vaccination (BNT162b2), blood samples were obtained from healthy, non-smoking individuals (relatively older male group, n=6 vs. younger female group, n=23), who did not have systemic diseases that required continuous treatment. Washed platelets were prepared and subjected to a fluorometric determination of platelet polyP levels using 4’,6-diamidino-2-phenylindole. The side effects of vaccination were recorded as scores.

**Results:** Compared with the male group, platelet polyP levels decreased in the relatively younger female group after the initial dose, while the side effect score increased in the female group after the second dose. Moderate correlation coefficients were observed between the reduction in polyP levels and the side effect scores or the original polyP levels.

**Conclusions:** Despite being a pilot study using a small sample size, this study suggests the possibility that platelet polyP may suppress the side effects induced by the mRNA vaccines after the initial dose, but not the second dose, in relatively young female subjects who generally have high immune responsiveness.

**Essentials:**

- The COVID-19 mRNA vaccines (BNT162b2) reduced platelet polyP levels after the initial dose, but not after the 2^nd^ dose, in relatively younger female subjects.
- Relatively older male subjects did not respond to the vaccination by reducing platelet polyP.
- These findings suggest that platelets release polyP to suppress vaccine-induced reactions, for example, inflammation, which is usually recognized as a side effect.
- However, such suppression could be observed in subjects with higher immune responses, generally in relatively younger female subjects.

## 1 INTRODUCTION

Platelets are known to be involved in hemostasis, thrombosis, and wound healing. However, recent studies have often highlighted their distinct functions in inflammation. Upon infection, platelets first bind to infectious pathogens, such as bacteria and viruses, secrete various immunoregulatory cytokines, and express receptors for mediating various immune effects and regulatory functions.^1^ Platelets regulate macrophage functions, regulatory T cells, and secrete pro-resolving mediators.^2^ However, their regulatory functions are not simple: both proinflammatory and anti-inflammatory regulations are included and modulated by the degree and the phase of injury and inflammation (for more details, see “4 Discussion section”).

The novel severe acute respiratory syndrome coronavirus 2 (SARS-Cov-2), which is the causative agent of the current pandemic of coronavirus disease-2019 (COVID-19) and which poses a great threat to the world, comprises the major components of a coronavirus, such as envelop, membrane, and spike proteins.^3^ The spike protein recognizes and binds the host cell surface receptor, angiotensin-converting enzyme 2 (ACE2), and aids the entry of SARS-Cov-2 into host cells^3^, including platelets, and probably similar to influenza virus.^1, 4, 5^ Initially, coronavirus infection elevates fibrinogen and D-dimer levels to evoke systemic hypercoagulability and frequent venous thromboembolic events.^6^ This could be considered as a severe proinflammatory state, which consequently induces pro-coagulopathy through endothelial activation/damage.^6^ As anticipated, the number of circulating activated platelets are found to increase in COVID-19 patients.^6-9^ This suggests that platelets contribute to COVID-19 severity,^7^ although the role of platelets in the pathogenesis of COVID-19 is not yet clarified.

There are several major types of COVID-19 mRNA vaccines available in Japan. Initially, the Pfizer-BioNTech vaccine (BNT162b2) was provided to medical staff as a priority and then to older residents. The Moderna vaccine (mRNA-1273) is the second major vaccine and is widely available to all applicants. Both vaccines are designed to induce the production of the spike protein of SARS-Cov-2 and then that of the spike protein-specific neutralizing antibodies associated with protective immunity.^10, 11^ Because these vaccines mimic the initial phase of reaction to SARS-Cov-2 infection, these agents also possibly cause various “side effects” that are originally recognized as symptoms of COVID-19.^12^ Thus, it is possible that the vaccines stimulate platelet activation and degranulation, in which polyP must be included, to modify inflammation and immunological reactions.

Among platelet regulatory molecules, polyphosphates (polyP) have recently received increasing attention not only as coagulation factors but also as potent modulators of inflammation. PolyP, a linear polymer of orthophosphate linked by high-energy phosphoanhydride bonds, is stored mainly in platelet dense granules and released upon activation.^13, 14^ PolyP modulates inflammation as a proinflammatory factor and an anti-inflammatory factor by triggering bradykinin release and inhibiting complement activation, respectively.^15, 16^ Thus, it can be anticipated that platelets activated by infectious pathogens release proinflammatory and anti-inflammatory biomolecules, including polyP, thereby exacerbating (or suppressing) inflammation.

To test this hypothesis, we first examined the effects of coronavirus vaccines on platelet polyP levels. Due to the lack of specific probes, it is difficult to accurately quantify platelet polyP levels in an efficient manner. In previous studies,^17, 18^ we optimized the previous methods, especially those related to sample preparation and the parameter of light wavelengths in fluorescent determination, for quantification and visualization of platelet polyP using DAPI. Although not completely excluding the possible count of DAPI-bound, non-polyP substances, this method enables rapid quantification of polyP levels without sacrificing recovery or reproducibility.

## 2 METHODS

### 2.1 Preparation of platelet suspension

The vaccination with mRNA vaccines, BNT162b2 and mRNA-1273, was controlled by the Japanese government and local governments. In principle, the residents were categorized into several groups depending on various factors, such as their age, severity of illness, area of residence, type of occupation, and scheduled for vaccination without or upon their requests and informed when they will be vaccinated.

Regarding to BNT162b2, healthy Japanese subjects [male: n=6 (age: 47−60) or 8 (age: 47−72), female: n=14 (age: 20−57) or 23 (age: 21−66)] who were not receiving continuous medical treatment participated in this study. The age distributions of the sample groups are shown in Figure 1. The female groups (initial dose=36.3±15.0, second dose=36.3±12.3) were significantly younger in average than the male groups (initial dose=56.8±8.8, second dose=56.0±7.8) in both cases.

**FIGURE 1.**
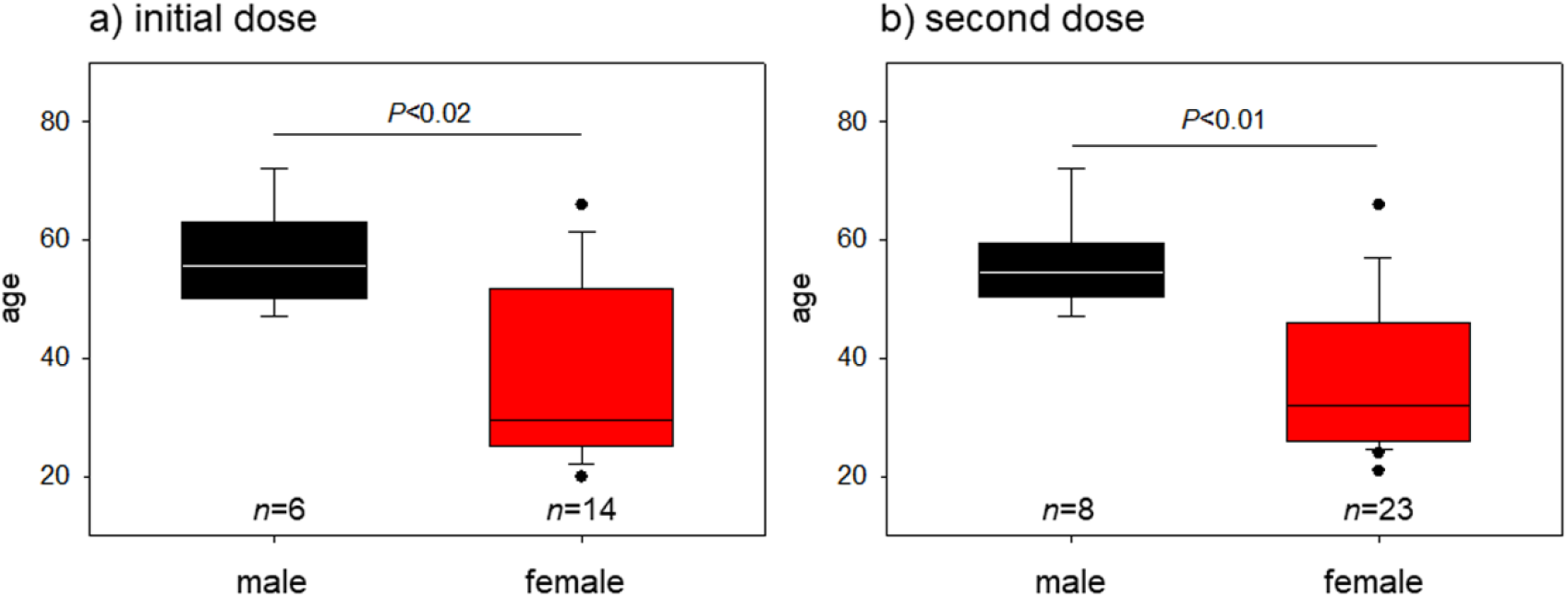
Age distribution of male and female groups of (a) the post-initial and (b) the post-second vaccination in the case of BNT162b2. The black and red columns represent male and female groups, respectively. The block dots represent outliers.

When the subjects were available during daytime on weekdays after appropriate intervals (initial dose: 4-10 days, median=8 days, second dose: 4-15 days, median=7 days), blood withdrawal was performed by venipuncture of the median cubital vein using 21G wing needles (NIPRO, Osaka, Japan). Blood was collected into vacutainer A-formulation of acid-citrate-dextrose (ACD-A) glass tubes (BD, Franklin Lakes, NJ, USA). Blood samples were subjected to blood cell count using an automated hematology analyzer (pocH-100i V Diff; Sysmex, Kobe, Japan) and intermittently rotated at 22–25 °C until use.

Prior to centrifugation, an excess amount of ACD-A (0.5 mL) (Terumo, Tokyo, Japan) was added to each blood sample and incubated for 10 min to prevent the unexpected coagulation that is often observed in female samples.^18^ Platelets were collected by the dual-spin protocol of pure-PRP preparation (1^st^ soft spin: 472× g for 10 min, 2^nd^ hard spin: 578× g for 5 min). The resulting platelet pellets were gently suspended in PBS, incubated for 15 min, and fixed with ThromboFix Platelet Stabilizer (Beckman-Coulter Life Sciences, Indianapolis, IN, USA). The fixed samples were stored at 4 °C for at least 24 h prior to polyP quantification.

The study design and consent forms for all procedures (project identification code: 2019–0423) were approved by the Ethics Committee for Human Participants at the Niigata University School of Medicine (Niigata, Japan) and complied with the Helsinki Declaration of 1964, as revised in 2013.

### 2.2 Fluorometric measurement of platelet polyphosphate using DAPI

After storage at 4 °C for 24 h, the fixed platelets were centrifuged at 578 × g for 5 min, and the supernatants were carefully aspirated as much as possible. The resulting platelet pellets were gently suspended in Milli-Q water. Platelets were counted using the automated hematology analyzer and adjusted to the appropriate range (<4.5×10^7^/mL) for polyP quantification. 4’,6-Diamidino-2-phenylindole (DAPI) (Dojindo, Kumamoto, Japan) was added to each tube at a concentration of 4 μg/mL and incubated with DAPI for 30 min at 22–25 °C. Fluorescence intensity was measured using a fluorometer (FC-1; Tokai Optical Co., Ltd., Okazaki, Japan) with excitation and emission wavelengths of 425 and 525 nm, respectively.^18^

The polyP ratio was calculated in each individual and totaled for statistical analysis.

### 2.3 Determination of mitochondria activity using cell-counting kit

The number of platelets suspended in PBS were adjusted to the density range of 2.5−5.0×10^7^/100 μL, mixed with 10 μL of cell-counting kit-8 (Dojindo), and incubated for 1 h at 22−25°C. After centrifugation (578 × g, 5 min), the supernatants were subjected to spectrophotometric determination at 450 nm (SmartSpec Plus; Bio-Rad, Hercules, CA, USA).

### 2.4 Evaluation of side effects based on individual self-declaration

The side effects are listed in Table 1. The perception of vaccine side effects was examined by asking the subjects, “Have you had any of these side effects? Please score each side effect on a scale where 0=not experienced, 1=experienced, and 2=experienced strongly (more important or severe for you). If you took painkillers, please score 1. Also, please mention the number of days the side effects lasted and the frequency of taking medicines against them.”

**TABLE 1.** List of vaccine-induced side effects and points given individual symptoms.

| **Systemic side effects** | **Point (a)** | **Running days or frequency of taking medicine (b)** | **Score: (a) × (b)** |
| --- | --- | --- | --- |
| fever (temp $\geq 37.5^{\circ}\text{C}$) | 2 | | |
| slight fever (temp $< 37.5^{\circ}\text{C}$ but $> 37.2^{\circ}\text{C}$) | 1 | | |
| fatigue | 1 |  |  |
| headache | 1 |  |  |
| others (chills, arthralgia, myalgia, nausea, etc.) | 1 |  |  |
| **Local side effects** |  |  |  |
| pain | 1 |  |  |
| redness | 1 |  |  |
| swelling | 1 |  |  |
| others (tenderness, itch, warmth, bruising, etc.) | 1 |  |  |
| **Allergic reaction** |  |  |  |
| rash | 1 |  |  |
| skin burning | 1 |  |  |
| red welts on face and lips | 1 |  |  |
| **Analgesics dose** |  |  |  |
| type of analgesics* | 1 |  |  |

*Note:* This score table was prepared by declaration of each subject.
\*Analgesics include nonsteroidal anti-inflammatory drugs (NSAIDs) and acetaminophen.

### 2.5 Statistical analysis

Each quantification was performed in triplicate. Data are expressed as the mean ± SD in the text and plotted in box plots and scatter plots. The Mann-Whitney U test was performed to compare the two groups (SigmaPlot 13.0; Systat Software, Inc., San Jose, CA, USA). Data for the apparent reduction of platelet polyP levels (<0.9) and the apparent side effect scores (≥10) in individual data were statistically analyzed using Fisher’s exact test. Differences were considered statistically significant at *P*<0.05.

Linear regression analysis and the calculation of correlation coefficient values were performed using SigmaPlot. Absolute R values ranging from 0.6 to 0.79 and 0.4 to 0.59 were considered a “strong” and “moderate” correlation, respectively. R values of 0.2 to 0.39 and lower than 0.2 indicated a “weak” and “very weak” correlation, respectively.

## 3 RESULTS

Ratios of polyP levels in post-vaccination (BNT162b2) to polyP levels in prior-vaccination, that is, vaccine-induced reduction of polyP levels, are shown in Figure 2a. There were no significant differences between the male and female groups in cases of either the initial dose (1.104±0.290 vs. 0.904±0.331) or the second dose (1.028±0.204 vs. 0.892±0.206). Scores for the side effects in the post-vaccination period are shown in Figure 2b. The scores were generally lower in male groups than in female groups; however, a statistical difference (*P*<0.02) was found only in the case of the second dose (2.88±2.42 vs. 8.22±6.2) (c.f., initial dose: 1.67±1.21 vs. 3.57±2.53).

**FIGURE 2.**
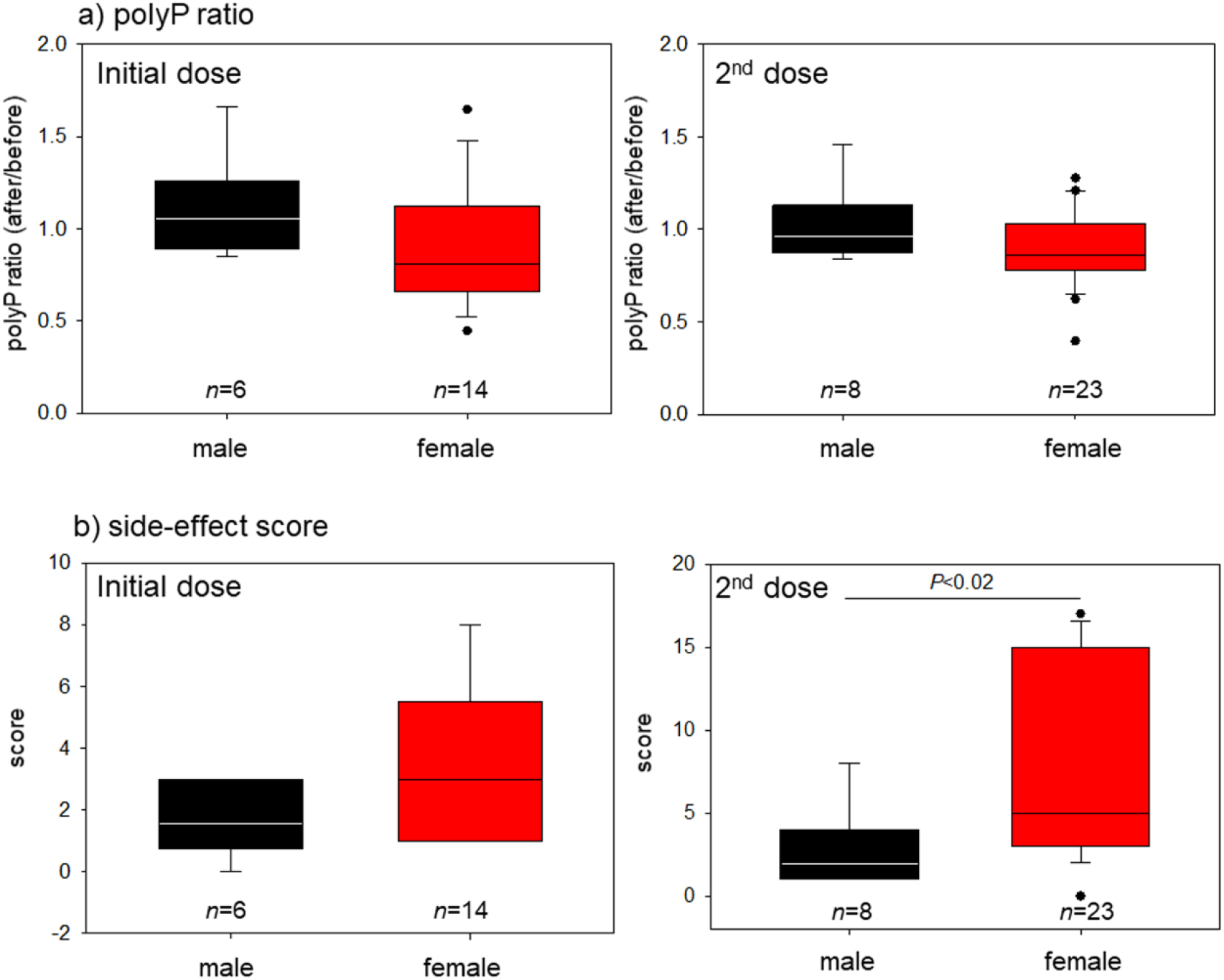
(a) polyP ratio and (b) side effect score distributions of the post-initial and the post-second vaccination to polyP levels in the post-vaccination in the case of BNT162b2. The black and red columns represent male and female groups, respectively. The block dots represent outliers.

To confirm the statistical difference, the data were analyzed again using Fisher’s exact test. A statistical difference (*P*=0.042) was found in the polyP ratios after the initial dose (male vs. female) (Table 2). On the other hand, no significant difference was found in the scores after the second dose (male vs. female; *P*=0.068), in which a statistical difference was found by the Mann-Whitney U test (Table 3).

**TABLE 2.** Number of subjects showing apparent reduction of platelet polyP by BNT162b2.

| Initial dose | Total number of subjects | polyP reduction <0.9 (%) | *P* value (Fisher's exact test) |
| --- | --- | --- | --- |
| male | 6 | 0 (0.0) | 0.042 |
| female | 14 | 8 (57.1) |  |
| Second dose |  |  |  |
| male | 8 | 3 (37.5) | 0.412 |
| female | 23 | 14 (60.9) |  |

**TABLE 3.** Number of subjects showing apparent exacerbation of vaccine side effect scores in the case of BNT162b2.

| Initial dose | Total number of subjects | polyP reduction ≥10 (%) | *P* value (Fisher's exact test) |
| --- | --- | --- | --- |
| male | 6 | 0 (0.0) | — |
| female | 14 | 0 (0.0) |  |
| Second dose |  |  |  |
| male | 8 | 0 (0.0) | 0.068 |
| female | 23 | 9 (39.1) |  |

**TABLE 4.**
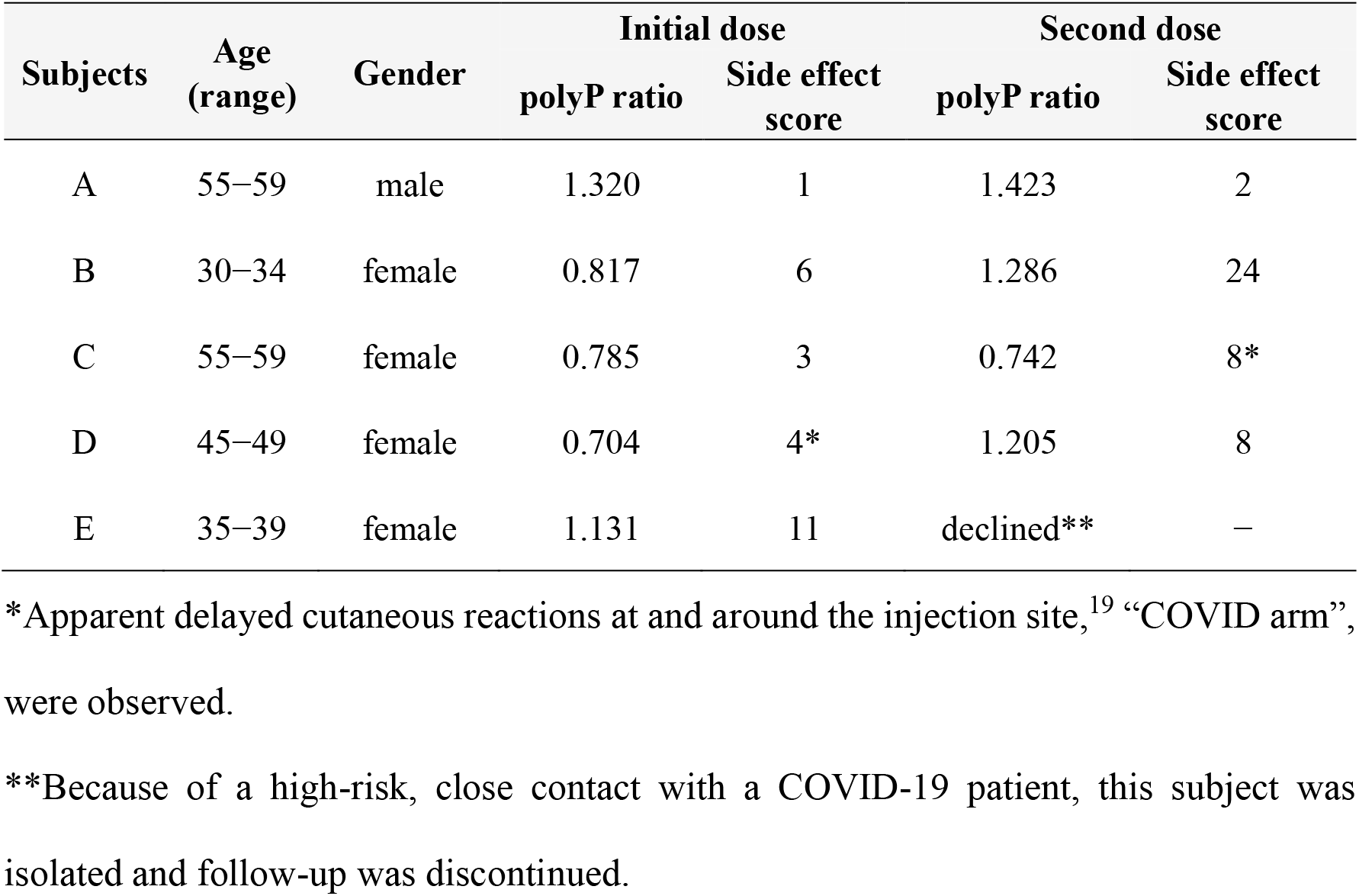
polyP ratios and side effect scores in the case of mRNA-1273.

Scatter plots between polyP ratios (after vaccination/before the initial vaccination) and side effect scores for BNT162b2 are shown in Figure 3. In the male groups, the coefficient correlations (R) were 0.279 and −0.115 for the initial and the second doses, respectively. In contrast, although it was very weak (R=0.142) for the second dose, a moderate correlation (R=0.556) was found for the initial dose.

**FIGURE 3.**
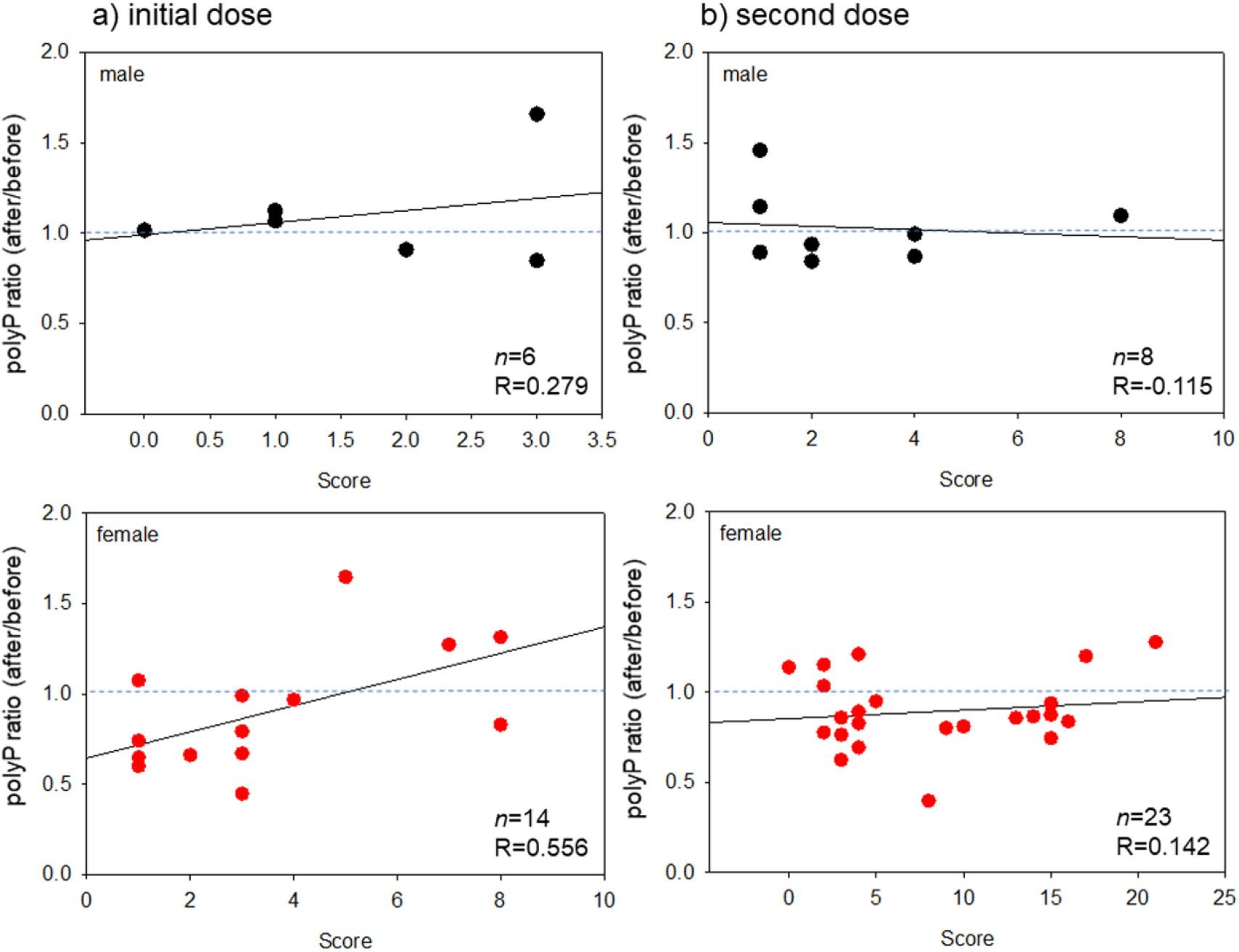
Scatter plots between polyP ratio (after the vaccination / before the initial vaccination) and side effect scores in (a) the post-initial and (b) the post-second vaccination in the case of BNT162b2. The black and red symbols represent male and female groups, respectively.

Scatter plots between polyP ratios (after vaccination/before the initial vaccination) and age in terms of BNT162b2 are shown in Figure 4. In the male groups, the correlation coefficients were weak (R=0.217) and very weak (R=0.143) for the initial and second doses, respectively. In the female groups, the correlations were negative, but very weak (R=−0.168) and weak (R=−0.224) for the initial and second doses, respectively.

**FIGURE 4.**
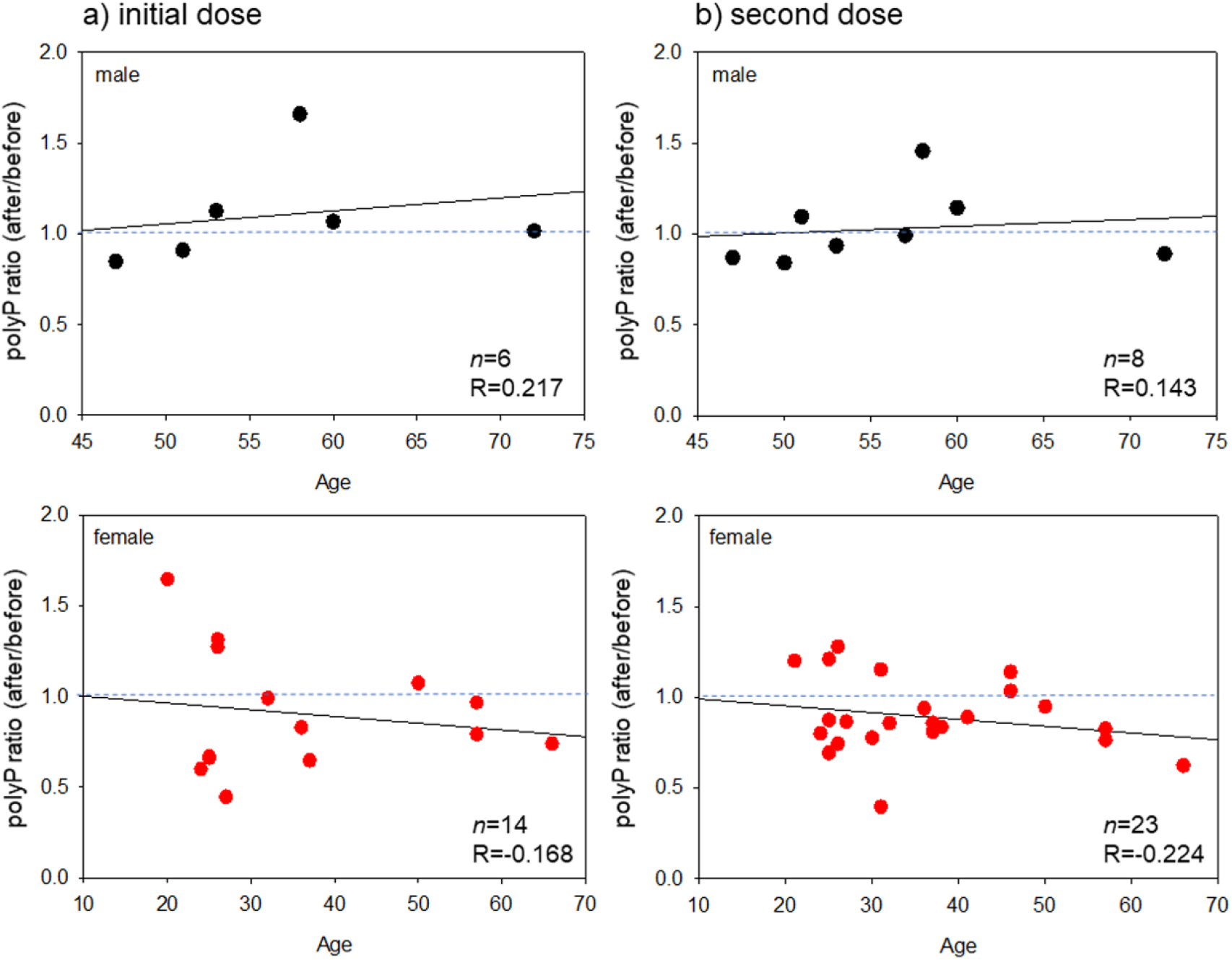
Scatter plots between polyP ratios (after the vaccination / before the initial vaccination) and age in (a) the post-initial and (b) the post-second vaccination in the case of BNT162b2. The black and red symbols represent male and female groups, respectively.

Scatter plots between the WST-8 reduction and polyP levels and the polyP ratio and original polyP levels in terms of BNT162b2 are shown in Figure 5. In the group containing both male and female subjects, a strong correlation (R=0.651) was found between NADH activity, which reflects general mitochondrial activity and platelet polyP levels (Fig. 5a). In the female group after the initial dose, a very strong negative correlation (R=−0.774) was found between the polyP ratio and the original platelet polyP levels (Fig. 5b).

**FIGURE 5.**
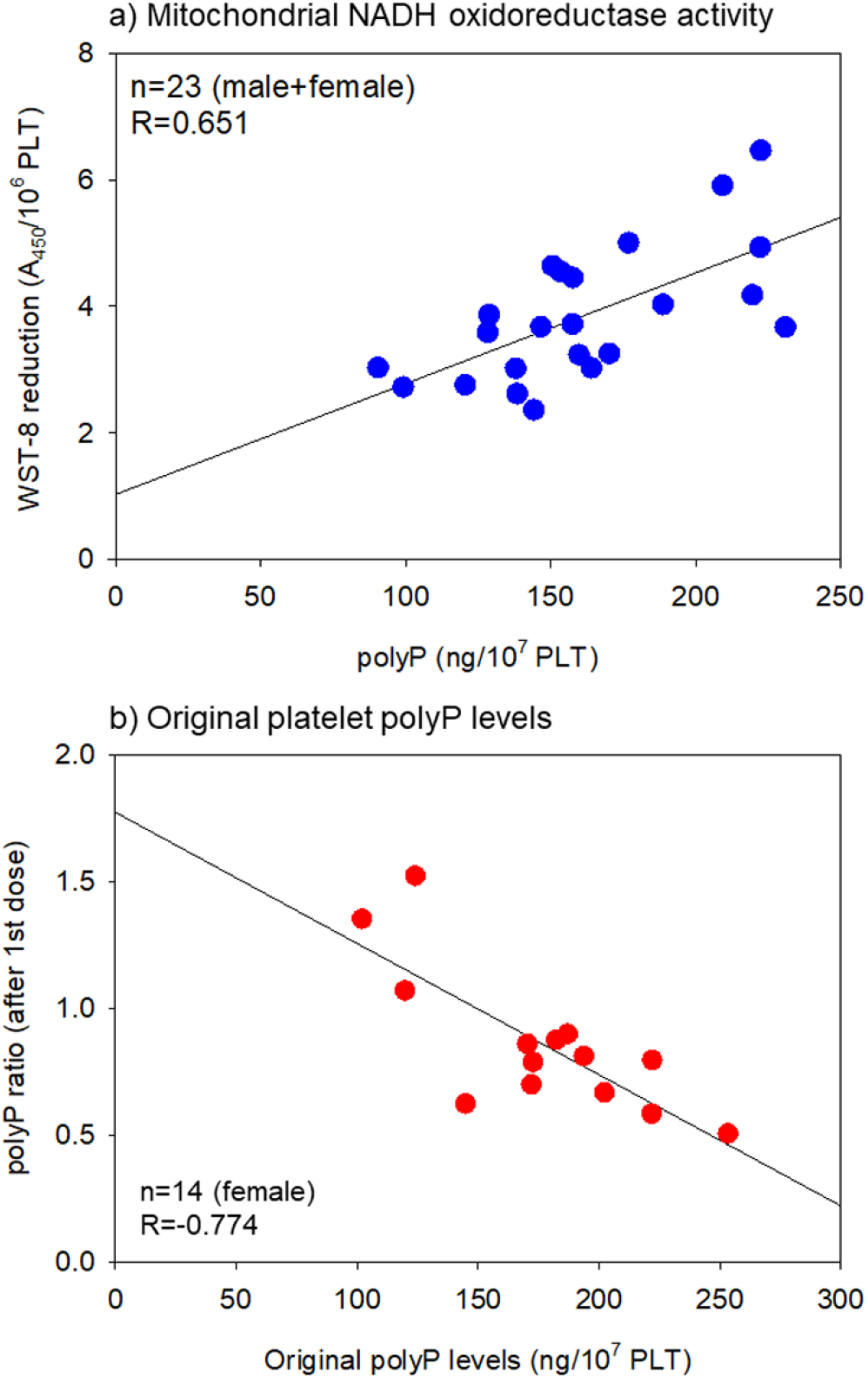
Scatter plots between (a) WST-8 reduction and polyP levels and (b) polyP ratio and original polyP levels in the post-initial vaccination in the case of BNT162b2. The blue and red symbols represent ‘male + female’ and female groups, respectively.

The effects of the other mRNA vaccine, mRNA-1273, were examined in a small number of subjects only due to its limited supply during our investigation. A substantial reduction in platelet polyP levels was observed in three female subjects (out of four) for the post-initial, whereas the polyP levels increased in three subjects (out of four) in the post-second doses. The oldest female subject was exceptional and showed both a low side effect score and COVID arm,^19^ both of which are usually observed after the initial dose. A drastic increase in the side effect score after the second dose was observed in only the youngest female subject (out of three).

## 4 DISCUSSION

### 4.1 Main findings of this study

Prior to designing this study, we hypothesized that if a micro-thrombosis is formed in the peripheral tissue, most probably around the site of vaccine injection, then platelet polyP is released (and intra-platelet polyP levels thereafter decrease) to function as a coagulation factor. However, no symptoms indicating local thrombocytopenia or purpura were observed on visual inspection. Although severe side effects, mainly represented by severe fever, headache, and pain at the injection site, were reported in several female subjects after the second dose, it was speculated that COVID-19 vaccination may not be able to induce platelet-dependent microthrombus formation or internal bleeding.

Interestingly, the vaccination of BNT162b2 significantly reduced platelet polyP levels after the initial dose, while the side effects seemed to be suppressed simultaneously in the younger female group. At present, we have no evidence to suggest how these phenomena are related, that is, a causal relationship or correlation, and further investigation is needed to clarify the involvement of polyP. However, a hypothesis to be considered for further investigation is that the spike protein produced by the mRNA vaccines induces inflammation, which is generally more severe in younger female individuals,^20^ and that the platelets activated by this inflammation release polyP along with other anti-inflammatory molecules to suppress the inflammation by a “negative feed-back loop.”

The dysfunction of this loop system after the second dose of BNT162b2 may be due to platelet status. Platelets contain various immunoreceptors and continuously scan for areas of injury or inflammation as versatile patrollers.^21, 22^ When platelets recognize intravascular pathogens, they directly limit pathogen growth and indirectly ensure pathogen clearance through the activation of immune cells.^2, 21, 23^ Thus, once the platelet signals are received and taken over by immune cells, inflammation may not be regulated by platelets. However, inflammation becomes more severe, and platelets may become dysfunctional. This scenario could be explained by a “chicken or egg” paradox-like essence introduced by Portier and Campbell that aberrant platelet activation, which could be caused by continuous moderate (or severe) inflammation, can lead to predominantly inflammatory and thrombotic events.^21^

### 4.2 Factors regulating platelet polyP metabolism

The abovementioned explanation assumes that the reduction of platelet polyP is due to possible polyP release from activated platelets. Before discussing other possible explanations, polyP metabolism in platelets should be confirmed. It is generally accepted that polyP is synthesized from ATP or GTP and degraded by cellular enzymes in bacteria.^15, 24, 25^ In eukaryotes, the mechanism is not yet fully understood; however, polyP is thought to be synthesized and degraded by the same or a similar mechanism. Inorganic phosphate (Pi) is incorporated by a specific transporter into mammalian cells and used for polyP synthesis in mitochondria non-enzymatically.^13^ Although polyP is widely distributed in the cytoplasm and organelles, it is the most condensed and stored in dense granules in platelets and released upon activation.^13^ Thus, it is generally accepted that platelet polyP levels are regulated by three major factors, that is, activation levels, mitochondrial activity, and extracellular Pi levels.

Since platelets of patients with sepsis or cardiac shock similarly have lower mitochondrial nicotinamide adenine dinucleotide dehydrogenase (NADH),^26^ in this study, we evaluated the activity of NADH using WST-8, a tetrazolium salt that is reduced to soluble formazan^27^ and found a moderate correlation between the activity of this enzyme and polyP levels. Serum phosphate levels were not examined in this study, but we did not receive any declaration of hyperphosphatemia or hypophosphatemia from individual participants. Taken together, it is most likely that the basal polyP levels depend on mitochondrial activity and influence the polyP reduction induced by platelet activation. Considering that platelet activation is a highly energy-dependent process,^28^ it is plausible that platelets with higher mitochondrial activity show higher immune responses to release polyP, resulting in polyP reduction.

### 4.3 Platelet activation by COVID-19 vaccines

In clinical observations, SARS-Cov-2 infection initially produces inflammation, platelet activation, and prominent elevation of fibrinogen and D-dimer.^6, 7, 9^ Subsequently, COVID-19 leads to various levels of thrombotic events and thrombocytopenia.^6^ mRNA vaccines are designed to temporarily provide spike protein at lower levels than those observed in SARS-Cov-2 infection or in DNA vaccines. Thus, even though the initial events induced by SARS-Cov-2 can be reproduced by the mRNA vaccines, theoretically, severe symptoms observed in COVID-19 patients cannot be produced. However, in reality, due to individual differences in immune activity, vaccines sometimes induce thrombotic thrombocytopenia, which is called VITT.^29, 30^ To date, VITT incidence has been recognized to be much lower in the case of mRNA vaccines than in DNA vaccines, such as ChAdOx1 nCov-19 and Ad26.COV2.5, but VITT has been reported in various groups and countries.^20^

Recently, the mechanism of VITT, mainly represented by heparin-induced thrombocytopenia (HIT), has been proposed.^31^ Platelet factor 4 (PF4) released from activated platelets binds blood heparin to form the complex (PF4-heparin), which binds to IgG antibodies and then binds to platelet FcγRIIa receptors to further activate platelets to induce thrombosis. Interestingly, the authors also mentioned the possibility that polyanions, such as polyP, may bind to PF4 to form the PF4-polyP complex and induce thrombosis. This possibility and the subsequent biological reactions should be further studied.

### 4.4 Possible platelet activation and thrombosis by polyP

In the earlier subsection, we described the possible anti-inflammatory effects of polyP, which are thought to be mediated through the suppression of the complement system.^32, 33^ However, the “conflicting” possibility that polyP has the potential to induce thrombosis should be noted. Surprisingly, although the chain length of polyP may be a matter for further investigation, it has been reported that polyanions like polyP can form a complex with PF4 and that this complex activates platelets as does the PF4-heparin complex.^31, 34^ Platelets also release anti-inflammatory cytokines, such as IL-4 and IL-10, upon activation,^1^ and thus platelet-rich plasma (PRP) has been clinically utilized as an anti-inflammatory agent or a pain-releasing agent.^35^ At present, it is difficult to predict the risk of PF4-polyP complex-induced thrombosis by the “autocrine” loop or by exposure to bacterial polyP after vaccination.

By applying this possibility to the findings of this study, we interpreted that the biological significance of these conflicting functions is that the unexpected aberrant activation and destruction of platelets and the uncontrolled exacerbation of inflammation and the immune system may be protected by the dual functions of platelets and polyP. Alternatively, especially in the younger female subjects who generally have higher immune activity, when exposed to the pathogens or maintained under inflammatory conditions for a long time (the interval between the initial and second dose in this study ≈3 weeks), platelets may lose the ability to detect pathogens and allow the exacerbation of inflammation, which can be recognized as side effects.

### 4.5 Limitations

In Japan, the vaccination of medical staff is strictly controlled and scheduled. In addition, our limited capacity to handle samples made it difficult to secure sufficient donor numbers. As a result, we could not expand the sample size to what we originally calculated. Thus, further investigation is needed to reach a definitive conclusion; however, the present findings regarding gender and age differences seem to essentially be consistent with the previous reports of antibody formation^36-38^ and, as a pilot study, are worth further investigation.

## 5 CONCLUSIONS

These findings suggest that polyP released from activated platelets may be involved in the suppression of possible severe side effects after the initial dose in young female subjects. This could be a reason why the young female subjects complained of severe side effects after the second dose, but not after the initial dose. The higher immune response is thought to be caused by estrogen.^39^ In addition, controlling such high immune responses may require our bodies to have more potent breaking systems. PolyP could function as one such system, and in this case, polyP could also be considered as a marker of the general activity of the immune response.

## Data Availability

The datasets used and/or analyzed during the current study are available from the corresponding author upon reasonable request.

## AUTHOR CONTRIBUTIONS

Conceptualization, T.U. and T.K.; methodology, T.K.; validation, T.Ts. and K.I.; formal analysis, T.W.; investigation, T.U., A.S., H.A., T.Ts., T.W., K.I., H.K., Y.K., and T.K.; data curation, H.A. and Y.K.; writing—original draft preparation, T.U., H.A., T.Ts., T.Ta., and T.K.; writing—review and editing, T.U., T.Ts., T.Ta., and T.K.; supervision, Y.K. and T.Ta.; project administration, T.W. and Y.K.; funding acquisition, T.K. All authors have read and agreed to the published version of the manuscript.

### Institutional Review Board Statement

The study design and consent forms for all procedures (project identification code: 2019– 0423) were approved by the Ethics Committee for Human Participants at the Niigata University School of Medicine (Niigata, Japan) and complied with the Helsinki Declaration of 1964, as revised in 2013.

### Informed Consent Statement

Informed consent was obtained from all subjects involved in the study.

